# Dyslipidemia in Young American Indians: Strong Heart Family Study

**DOI:** 10.1101/2023.06.13.23291360

**Authors:** Jessica A. Reese, Mary J. Roman, Jason F. Deen, Tauqeer Ali, Shelley A. Cole, Richard B. Devereux, Amanda M. Fretts, Wm. James Howard, Elisa T. Lee, Kimberly Malloy, Parmanand Singh, Jason G. Umans, Ying Zhang

## Abstract

**Background:** Although many studies on the association between dyslipidemia and cardiovascular disease (CVD) exist in older adults, data on the association among adolescents and young adults living with disproportionate burden of cardiometabolic disorders are scarce.

**Methods:** The Strong Heart Family Study (SHFS) is a multi-center, family-based, prospective cohort study of CVD in American Indians, including 12 communities in central Arizona, southwestern Oklahoma, and the Dakotas. We evaluated SHFS participants, 15-39 years old at the baseline examination in 2001-2003 (n=1,440). Lipids were measured after a 12-hour fast. We used carotid ultrasounds to detect plaque at baseline and follow-up in 2006-2009 (median follow-up=5.5 years). We identified incident CVD events through 2020 with a median follow-up of 18.5 years. We used shared frailty proportional hazards models to assess the association between dyslipidemia and subclinical or clinical CVD, while controlling for covariates.

**Results:** Baseline dyslipidemia prevalence was 55.2%, 73.6%, and 78.0% for participants 15-19, 20-29, and 30-39 years old, respectively. Approximately 2.8% had LDL-C ≥ 160 mg/dL, which is higher than the recommended threshold for lifestyle or medical interventions in young adults of 20-39 years old. During follow-up, 9.9% had incident plaque (109/1,104 plaque free participants with baseline and follow-up ultrasounds), 11.0% had plaque progression (128/1,165 with both baseline and follow-up ultrasounds), and 9.2% had incident CVD (130/1,416 CVD free participants at baseline). Plaque incidence and progression was higher in participants with total cholesterol ≥200 mg/dL, LDL-C ≥160 mg/dL, or non-HDL-C ≥130 mg/dL, while controlling for covariates. CVD risk was independently associated with LDL-C≥160 mg/dL.

**Conclusions:** Dyslipidemia is a modifiable risk factor that is associated with both subclinical and clinical CVD even among the younger population of American Indians who have unexpectedly high rates of significant CVD events. Therefore, this population is likely to benefit from a variety of evidence-based interventions including screening, educational, lifestyle and guideline directed medical therapy at an early age.

**Clinical Perspective:** *What is new?:*

- First report of lipid levels and dyslipidemia in American Indians under 40 years old, in a large, prospectively followed, multi-site cohort.
- The prevalence of dyslipidemia was 55.2%, 73.6%, and 78.0% for participants 15-19, 20-29, and 30-39 years old, respectively, which was more than twice as high than the general U.S. population in every age group.
- Abnormal lipid levels were independently associated with incident plaque, progression of plaque, and the hard clinical end points of cardiovascular disease morbidity and mortality.

*What are the clinical implications?:*

- Screening and management of dyslipidemia for the prevention of cardiovascular diseases are needed among adolescents and young adults who are predisposed to high risk of cardiometabolic disorders.
- Evidence-based interventions that include cholesterol health education, lifestyle modification, and guideline directed medical therapy may be used in preventing and managing dyslipidemia in the aforementioned population.

## Introduction

Dyslipidemia is an established risk factor for subclinical and clinical cardiovascular disease (CVD) in older adults.^1,2^ CVD is the leading cause of mortality in the United States (U.S.), accounting for approximately one third of all deaths.^3^ Through systematic surveillance by the Strong Heart Study (SHS), a study of CVD in American Indians, we have reported that CVD incidence and mortality continue to be higher in American Indians compared to other U.S. populations.^4^ Among a SHS population of adults aged 45-74 at baseline, high LDL-C^5^, high TG, and low HDL-C^1^ were associated with coronary heart disease (CHD) and stroke, despite recent advances in treatment of CVD and related risk factors^1,6^ Due to the association between dyslipidemia and CVD and the rising rates of CVD in older adults, lowering LDL-C levels with statin therapy along with adherence to a healthy lifestyle has become the primary target of reducing subclinical and clinical CVD.^1,7-9^ Intervention is even more important in people with diabetes, where aggressive LDL-C (≤70 mg/dL) and blood pressure control has improved atherosclerosis when compared to standard targets.^10^ While the beneficial evidence for lowering LDL-C has been established, several large studies have recently reported that drug interventions designed to increase HDL-C have not improved CVD outcomes when added to standard therapy.^11^ Therefore, more research is vital to investigate the association between dyslipidemia and CVD outcomes in young American Indians.^12,13^

Although numerous studies relating dyslipidemia to CVD exist in older adults, the data on adolescents and young adults are scarce. The prevalence of dyslipidemia in the entire population of U.S. adolescents is approximately 25%;^14^ and in adults aged 18-39, it is approximately 30%.^15^ However, the prevalence of dyslipidemia in American Indian adolescents and young adults has not been described.^16^ In addition, we have not investigated the potential associations between dyslipidemia and subclinical or clinical CVD in this younger population. To address these gaps in knowledge, we assessed the prevalence of dyslipidemia and examined its association with subclinical and clinical CVD among American Indians, ages 15 to 39 years, who participated in the baseline and follow-up examinations of the Strong Heart Family Study (SHFS).

## Methods

### Study population

The SHFS is a multi-center, family-based, prospective cohort study of CVD in American Indians.^16^ It includes 12 American Indian communities and tribes in Arizona, Oklahoma, and the Dakotas.^17^ For this present study, we included participants who were 15-39 years old at the baseline SHFS examination (n=1,440, conducted between 2001-2003) and had lipid measurements during the examination (n=1,424, 12 excluded for missing lipid measurements). For the outcome assessments, we further excluded participants with prevalent CVD, based on chart review (n=1,416, 12 excluded for prevalent CVD). We defined CVD as history of any one of the following diagnoses based on chart review: coronary heart disease (CHD) including myocardial infarction (MI), stroke, or congestive heart failure (CHF), the definitions of which have been described previously.^16,18^ Since we need ultrasound measurements at both baseline and follow-up to calculate plaque incidence and progression, we excluded participants from these analyses who did not have measurements at both timepoints (Figure 1). There were 13 participants with missing ultrasounds at baseline, 241 were missing at follow-up, and 3 were missing both baseline and follow-up ultrasounds (n=251 at baseline or follow-up). The SHFS protocols and procedures were approved by participating tribal communities, and IRBs of Indian Health Service and the participating institutions.^18^ We obtained informed consent from every participant.

**Figure 1.**
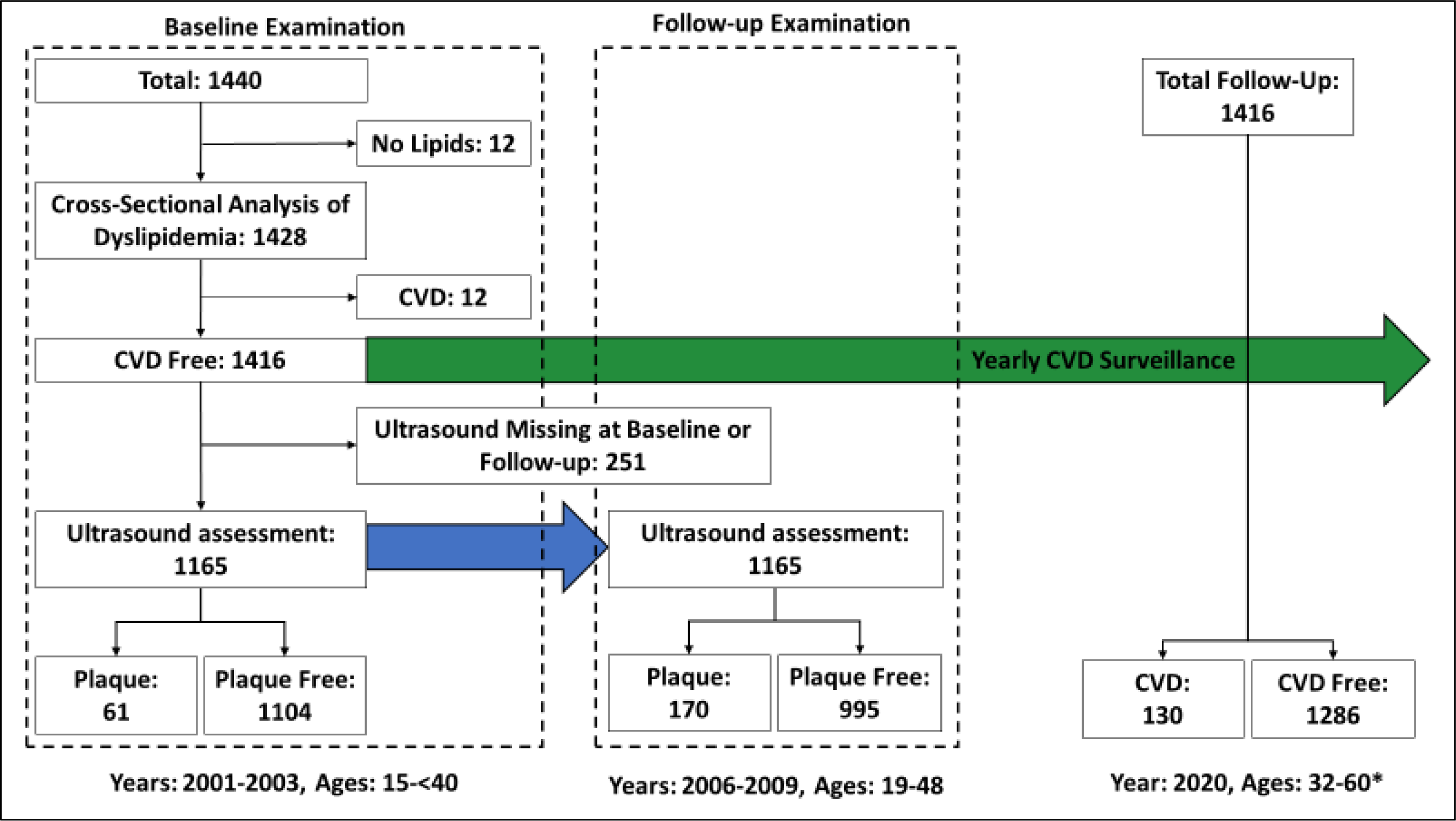
Sample sizes and length of follow-up for the Strong Heart Family study baseline assessments of American Indians ages 15 to <40. Of the 1,440 American Indians who met the age requirements, 12 has missing lipid assessments. We excluded these participants from the dyslipidemia prevalence analysis, leaving 1,428. Of the 1,428 with lipid assessment, 12 had cardiovascular disease (CVD) at baseline according to chart review. We excluded these participants from the incident CVD analysis, since they had prevalent CVD, leaving 1,416 with yearly CVD surveillance. Of the participants who were CVD free at baseline, 1,165 had ultrasounds at both baseline and follow-up. There were 13 participants with missing ultrasounds at baseline, 241 were missing at follow-up, and 3 were missing both baseline and follow-up ultrasounds, leaving 251 who were missing ultrasound measurements at baseline or follow-up. Of the 170 who had plaque at follow-up, 61 had plaque at baseline, leaving 109 with incident plaque. *This is the age range of surviving participants as of December 31, 2020. At the end of surveillance follow-up, 121 (8.5%) deaths had occurred. Of these 121 deaths, 30 (25%) were CVD related.

### Dyslipidemia assessment

To measure lipids, we drew blood after a 12-hour fast.^19,20^ Total cholesterol (TC) and triglycerides (TG) were directly measured and we estimated low density lipoprotein cholesterol (LDL-C) using the Friedewald equation, except for samples that had triglycerides > 400 mg/dL, where we directly measured LDL-C (n=35).^21,22^ High density lipoprotein cholesterol (HDL-C) was measured after precipitation of LDL-C and very low-density lipoprotein cholesterol (VLDL-C).^23^ We defined dyslipidemia as any of the following: 1. TC ≥200 mg/dL; 2. LDL-C ≥100 mg/dL; 3. HDL-C <40 mg/dL for men or HDL-C <50 mg/dL for women; 4. non-HDL-C ≥130 mg/dL; 5. TG ≥150 mg/dL; or 6. taking lipid lowering medication.^12,20^ We defined very high LDL-C as ≥160 mg/dL, which is the recommended threshold for medical along with lifestyle interventions in this population.^9^ Lipid lowering medication was based on the American Hospital Formulary Service Pharmacologic-therapeutic classification of 24:06.^24^

### Subclinical atherosclerosis assessment

During the baseline and follow-up (between 2006 and 2009) SHFS examinations, we performed carotid ultrasonography using a standardized protocol.^19,25,26^ We used Acuson 128 machines equipped with a 7.5-MHz imaging transducer to assess the left and right carotid arteries for presence of atherosclerotic plaque, defined as arterial wall thickening (focal protrusion) at least 50% greater than the surrounding wall.^19,27^ To evaluate the extent of the atherosclerosis, we calculated a plaque score by determining the number of segments of the left and right common carotid, bulb, internal carotid, and external carotid arteries containing plaque^19^. Plaques scores could range from 0 (no plaque) to 8 (plaque observed in all studied segments), and the larger the score, the greater the scope of vascular atherosclerosis. Centrally trained, research sonographers performed all ultrasound studies, and a cardiologist, blinded to the clinical characteristics of participants, interpreted the results. Quality control was performed for extreme values.^19,25-27^ We defined incident plaque as the presence of plaque at the follow-up examination among participants free of plaque at the baseline examination. We defined the progression of plaque as the number of plaques at follow-up that was higher than the number of plaques at baseline.

### Clinical CVD assessment

To determine the occurrence of fatal or non-fatal CVD events, trained medical record abstractors reviewed and abstracted events on all participants yearly from 2001 through 2020.^4,28^ Sources of data for physicians’ review include international classification of diseases codes, medical charts, autopsy reports, death certificates, and informant interviews.^28^ The morbidity and mortality review procedure and criteria of confirming morbid or fatal CVD events have been reported in detail previously.^17,28,29^ For fatal CVD events, two physicians on the SHS mortality review committee used standard criteria to independently review documents collected by abstractors.^17,28^ If they did not agree, a senior reviewer adjudicated the case. We defined fatal CVD events as death from the following: sudden death due to CHD, other CHD including MI, stroke, CHF, or other CVD, such as cardiomyopathy. SHS investigators have previously published the criteria used for the primary CVD death categories.^28,30,31^

### Covariate assessment

We included covariates reported to be associated with both subclinical and clinical CVD.^32-35^ We collected self-reported demographic and clinical characteristics. We asked participants to bring their medications to the physical examination and to recall (with assistance from an adult for minors) additional medication.^34^ We measured height, weight, and waist circumference during the physical examination. An increased waist circumference was defined as >100 cm for men or >90 cm for women.^36^ We measured blood pressure from the right arm three times while the participant was in the seated position after five minutes of rest, and used the average of the second and third blood pressure measurements for analysis.^18,34^ Hypertension was defined as systolic blood pressure (SBP) ≥ 140 mm Hg, diastolic blood pressure (DBP) ≥ 90 mm Hg, or taking antihypertensive medication.^34^ To measure fibrinogen, plasma glucose, and creatinine, we drew blood after a 12-hour fast.^19,20^ Diabetes was defined as a fasting plasma glucose level ≥126 mg/dL or use of anti-diabetes medication^18,37^ Since albuminuria is a significant predictor of CVD in this population, we included it as a covariate,^38,39^ defined as albumin-creatinine ratio ≥30 mg/g.^19,33^ Finally, we defined metabolic syndrome when at least three of five components for the syndrome were present: increased waist circumference (defined above), high TG, low HDL-C (defined above), high blood pressure (>130/85 mm Hg); or high fasting glucose (>100 mg/dL).^40^

## Statistical Analysis

We summarized the prevalence of dyslipidemia measures in three age groups: 15-19, 20-29, and 30-39. We compared demographic and CVD risk factors between participants with or without incident plaque, progression of plaque, or clinical CVD. For continuous, normally distributed variables, we used t-tests to determine if means differed between the groups. We used Wilcoxon rank sum tests for skewed continuous variables and chi-square tests for categorical variables.

Incidence rates of atherosclerotic plaque and clinical CVD were calculated using person-time, according to the status of baseline dyslipidemia. Similarly, we calculated incidence of CVD according to distinct categories of risk factors, including age group (divided at median of the study population), gender, current smoking, increased waist circumference, hypertension, diabetes mellitus, albuminuria, and metabolic syndrome. We used log-rank tests to determine if plaque incidence, plaque progression, or clinical CVD differed between the categories. In addition, we used Kaplan-Meier curves and log-rank tests to determine if the probabilities of incident plaque, progression of plaque, or clinical CVD differed between participants with or without baseline dyslipidemia.^41^

We assessed the relationship between dyslipidemia and subclinical/clinical CVD, adjusting for covariates, using a shared frailty model based on proportional hazards to account for the relatedness among participants.^42,43^ We used this method to calculate hazard ratios of associations between dyslipidemia and time to incident plaque, plaque progression, or CVD events. Each reported analysis met the assumption of proportional hazards. Initially, we evaluated the associations while controlling for age and sex. Then we added smoking, waist circumference, hypertension, diabetes, and albuminuria to the models. We then reduced the models, using backwards selection, by removing insignificant variables (p>0.05) from the full models.^44^ Interaction between covariates and dyslipidemia was evaluated by including appropriate cross-product terms in the model and no significant interactions were found.

Finally, we considered models separately for participants with and without baseline diabetes. We defined the time to event for each analysis as the time from baseline to the onset of plaque, plaque progression, clinical CVD onset, death of an individual regardless of cause, the date of last follow-up, or the end of the study period (December 31, 2020).^19^ We used a significance level of 0.05 for hypothesis tests and performed statistical analyses using SAS^©^ software, version 9.4 (2002-2012 by SAS Institute Inc., Cary, NC, USA.).

## Results

### Prevalence of dyslipidemia

At the baseline examination, 2001-2003, the median age was 26.8 years (range = 15-39 years) for those included in the dyslipidemia assessment (n=1,424). The prevalence of dyslipidemia was 55.2%, 73.6%, and 78.0% for participants 15-19, 20-29, and 30-39 years old, respectively (Table 1). The prevalence of LDL-C≥160 mg/dL was 2.8% (n=39). One participant 20-29 years old and 10 participants 30-39 years old were taking lipid lowering medications. Six participants were taking Atorvastatin, one was taking Lipitor, and four were taking Simvastatin (including the participant 20-29 years old).

**Table 1.** Prevalence of dyslipidemia measures according to age groups in American Indian adolescents and young adults from the Strong Heart Family Study.

| Variables | Mean (standard deviation) or percentage |  |  |  |
| --- | --- | --- | --- | --- |
|  | Total<br>n= 1,428 | Age 15-19<br>n=368 | Age 20-29<br>n=496 | Age 30-39<br>n=564 |
| Total Cholesterol (mg/dL) | 175.1 (38.4) | 153.1 (28.7) | 176.7 (34.2) | 188.0 (41.1) |
| High Total Cholesterol (% ≥ 200 mg/dL) | 22.7 | 7.1 | 24.2 | 31.6 |
| LDL-C (mg/dL) | 95.9 (28.8) | 82.1 (23.9) | 97.7 (27.8) | 103.4 (29.4) |
| High LDL-C (% ≥ 100 mg/dL) | 41.0 | 21.5 | 43.7 | 51.4 |
| Very High LDL-C (% ≥ 160 mg/dL) | 2.8 | 0.8 | 2.8 | 4.0 |
| HDL-C (mg/dL) | 50.2 (13.4) | 49.3 (12.7) | 50.0 (13.0) | 51.1 (14.1) |
| Low HDL-C (% < 40 for men, < 50 for women) | 39.9 | 41.4 | 39.3 | 39.4 |
| Non-HDL-C (mg/dL) | 124.2 (36.1) | 103.8 (29.9) | 126.8 (35.2) | 135.5 (35.1) |
| High Non-HDL-C (% ≥ 130) | 39.5 | 16.9 | 40.9 | 53.1 |
| Triglycerides (mg/dL)* | 119.0 (86.0-170.0) | 91.5 (72.0-128.0) | 120.0 (89.0-178.0) | 135.0 (99.0-188.5) |
| High Triglycerides (% ≥ 150) | 33.4 | 16.9 | 34.1 | 43.4 |
| Taking lipid lowering medication (% yes) | 1.8 | 0 | 0.5 | 3.1 |
| Dyslipidemia (% yes) | 70.6 | 55.2 | 73.6 | 78.0 |

\*median and interquartile range presented, LDL-C=low-density lipoprotein cholesterol, HDL-C=high-density lipoprotein cholesterol, non-HDL-C=non-high-density lipoprotein cholesterol, n=number.

### Association between dyslipidemia and atherosclerotic plaque

Of participants included in the assessment of subclinical atherosclerosis with ultrasound measurements at both baseline and at follow-up (n=1,165), 61 had baseline plaque and 1,104 were plaque free. Of the 1,104 plaque free participants at baseline, 109 (9.9%) developed plaque during follow-up. The plaque incidence rate during a median follow-up of 5.5 years was 15.6 per 1,000 person-years. Plaque progression included both incident plaque (progressing from no plaque to at least one plaque, n=109) and progressing from prevalent plaque at baseline to more affected segments at follow-up (n=19); therefore, the total number of participants with plaque progression was 128 (11.0%, 109 incident cases plus 19 with prevalent plaque that progressed).

Participants with incident plaque or plaque progression at the follow-up examination in 2006-2009, were more likely to be older, be a smoker, have prevalent hypertension, diabetes, or metabolic syndrome at baseline (Table 2). In addition, those with high TC, LDL-C, non-HDL-C, triglycerides, or dyslipidemia had significantly higher plaque incidence or progression (Figure 2). In the time-to-event analysis, participants with baseline dyslipidemia had a greater probability of developing incident plaque or plaque progression during follow-up (Figure 3).

**Figure 2.**
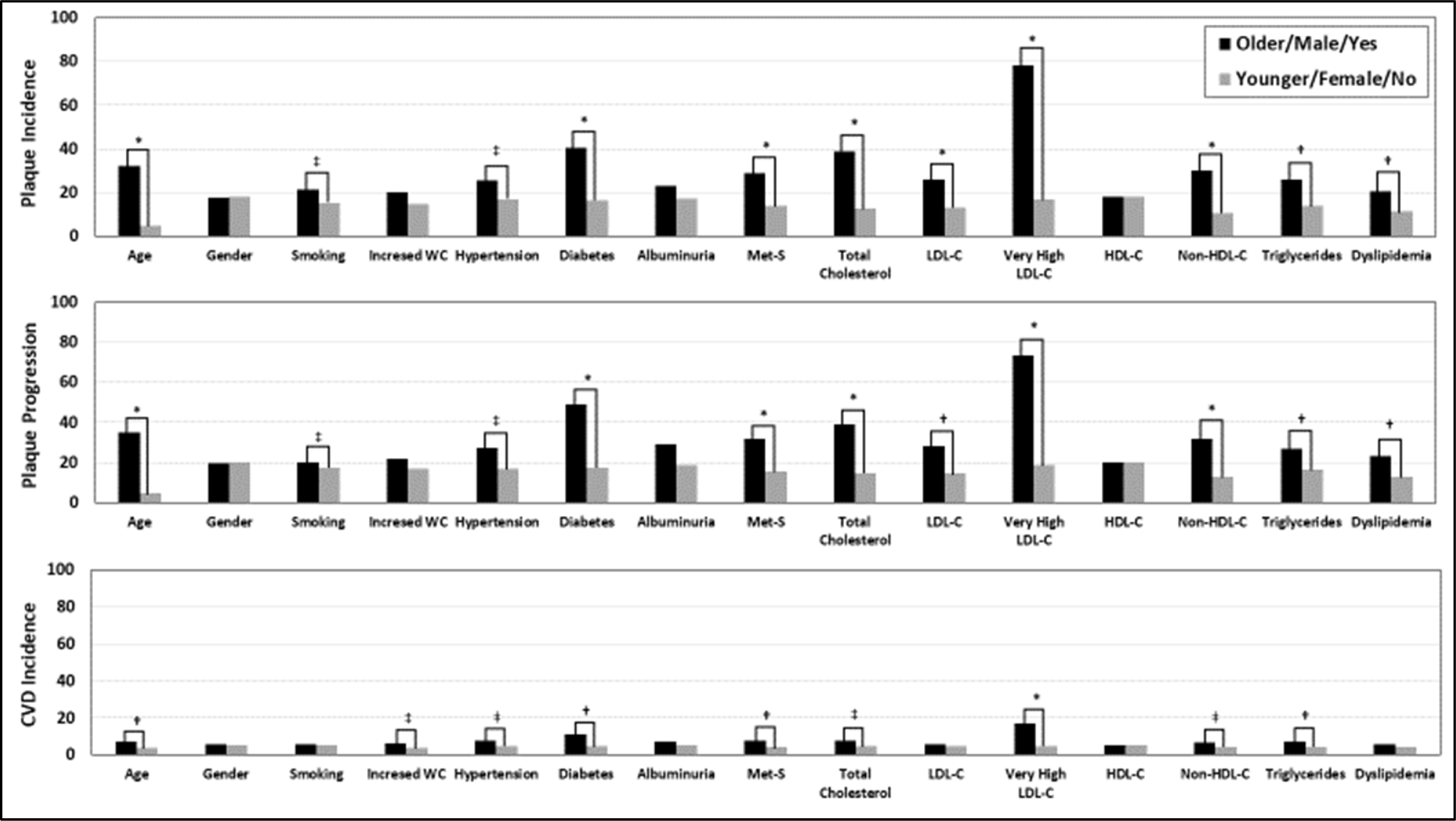
Plaque incidence, plaque progression, and cardiovascular disease (CVD) Incidence according to baseline demographic and cardiovascular disease risk factors in American Indian adolescents and young adults, ages 15 to <40, during a median of 5 years of follow-up for plaque outcomes and 19 years for CVD outcomes. Age is divided at median of the study population (26.8 years), increased waist circumference (WC) is defined as > 100 cm for men and >90 cm for women, hypertension is ≥ 140/90 mm Hg, diabetes is fasting plasma glucose level ≥126 mg/dL or the use anti-diabetes medication, albuminuria is an albumin-creatinine ratio ≥30 mg/g, metabolic syndrome (met-S) is when at least three of five components for the syndrome were present-increased WC, high triglycerides, low high density lipoprotein cholesterol (HDL-C), hypertension, or high fasting glucose (>100 mg/dL), total cholesterol is ≥200 mg/dL, low density lipoprotein cholesterol (LDL-C) is ≥100 mg/dL, very high LDL-C is ≥160 mg/dL, HDL-C is <40 mg/dL for men or <50 mg/dL for women, non-HDL-C is ≥130 mg/dL, triglycerides is ≥150 mg/dL, and dyslipidemia is the presence of any abnormal levels of total cholesterol, LDL-C, HDL-C, non-HDL-C, triglycerides, or taking lipid lowering medication. *p<0.0001, †p<0.01, ‡p<0.05, comparisons without these symbols were not statistically significant; p-values are from log-rank tests.

**Figure 3:**
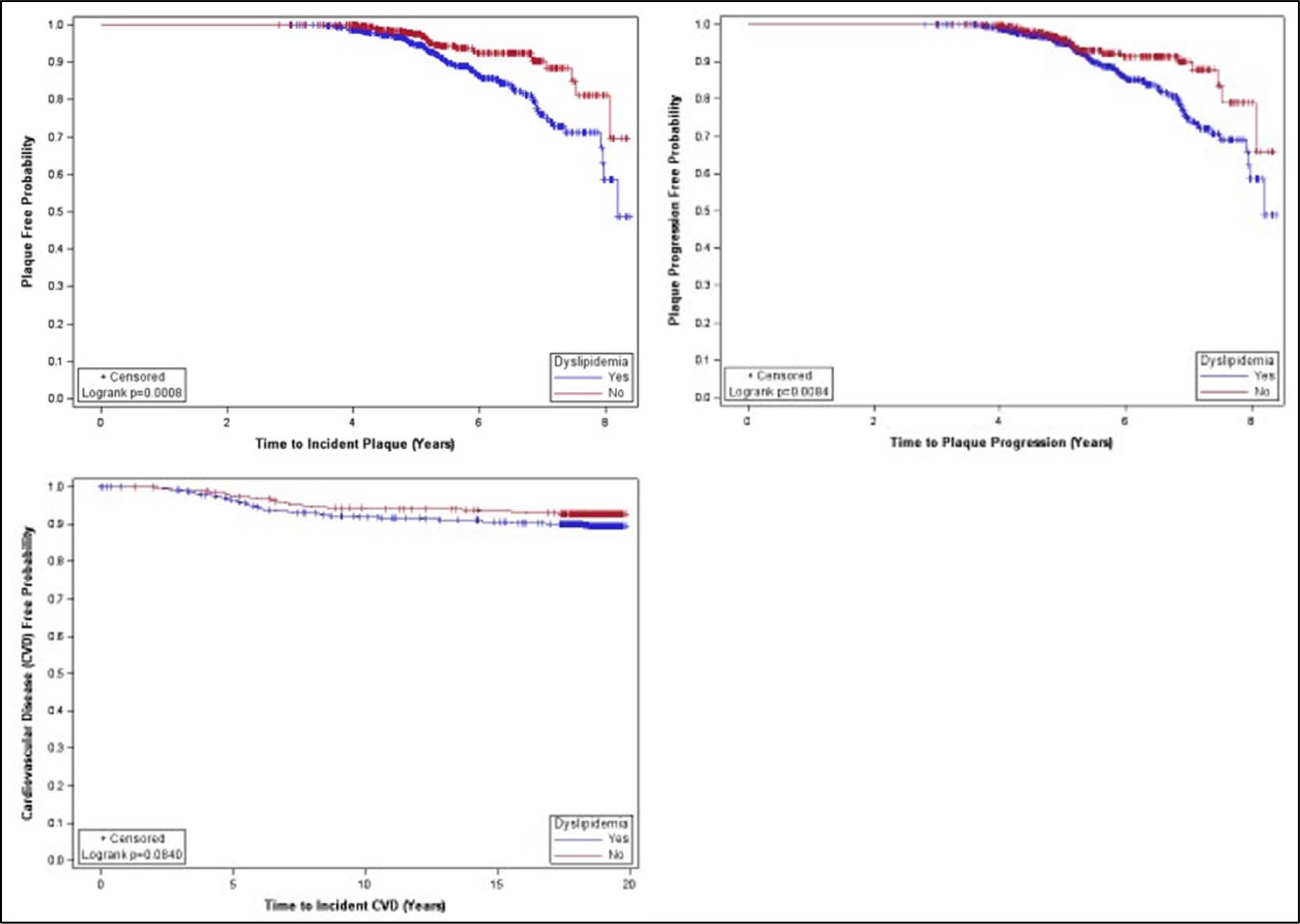
Plaque, plaque progression, and cardiovascular disease (CVD) free probability for American Indians, ages 15 to <40, with versus without dyslipidemia at baseline. Dyslipidemia is defined as any abnormal levels of total cholesterol (≥200 mg/dL), low density lipoprotein cholesterol (≥100 mg/dL), high density lipoprotein cholesterol (<40 mg/dL for men or <50 mg/dL for women), non-high density lipoprotein cholesterol (≥130 mg/dL), triglycerides (≥150 mg/dL), or taking lipid lowering medication. Blue and red lines represent the times to plaque development, plaque progression, or CVD for participants with or without baseline dyslipidemia, respectively. The plus symbols (+) represent censored participants.

**Table 2.**
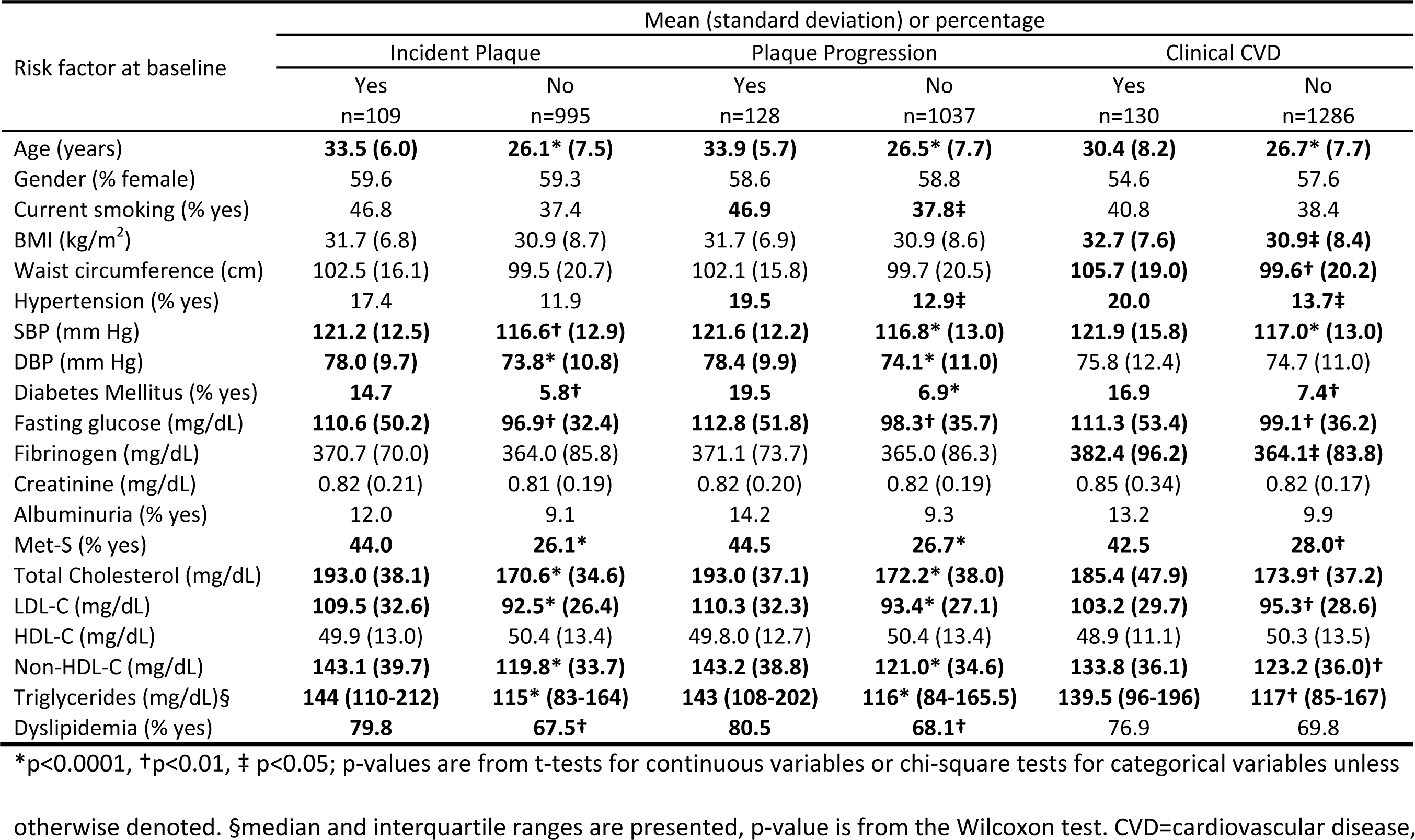
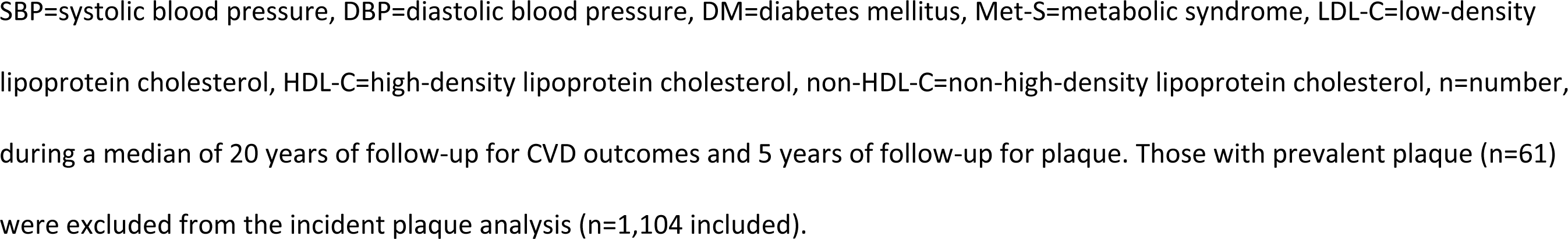
Comparison of baseline demographic and CVD risk factors in American Indian adolescents and young adults who were CVD free at baseline and subsequently developed incident plaque, plaque progression, and/or clinical cardiovascular disease.

In Table 3, we present the results of the multivariable shared frailty models of the association between different measures of dyslipidemia and incident plaque or plaque progression. The results were similar between the three separate models-age and sex adjusted, full model (adjusted for age, gender, smoking, waist circumference, hypertension, diabetes, and albuminuria), and reduced model. At an alpha of 0.05, age and diabetes remained in the reduced models. Although diabetes remained in the models, we did not observe any interactions with diabetes, and we did not observe statistically significant associations in the models stratified on diabetes. After adjusting for covariates, the risk of incident plaque or plaque progression was independently associated with elevated levels of TC, LDL-C, or non-HDL-C. This indicates that abnormal cholesterol levels are not only associated with the development of plaque but also the worsening of plaque.

**Table 3.** Multivariable Cox regression models of associations between measures of dyslipidemia and incident atherosclerotic plaque or cardiovascular disease in American Indian adolescents and young adults.*

| Measure of dyslipidemia | Hazard Ratios (95% Confidence Intervals) |  |  |
| --- | --- | --- | --- |
|  | Age and Sex adjusted model | Multivariable model† | Reduced model† |
| **Incident Atherosclerotic Plaque** |  |  |  |
| Total cholesterol (≥200 vs. <200 mg/dL) | **2.18 (1.45-3.27)** | **2.08 (1.37-3.17)** | **1.98 (1.31-2.99)** |
| LDL-C (≥100 vs. <100 mg/dL) | 1.26 (0.84-1.88) | 1.20 (0.80-1.81) | 1.20 (0.81-1.79) |
| LDL-C (≥160 vs. <160 mg/dL) | **3.30 (1.59-6.83)** | **3.49 (1.62-7.49)** | **2.97 (1.43-6.18)** |
| LDL-C (mg/dL, continuous) | **1.010 (1.004-1.017)** | **1.010 (1.003-1.017)** | **1.009 (1.002-1.015)** |
| HDL-C (<40 vs. ≥40 mg/dL for men and <50 vs. ≥50 mg/dL for women) | 1.03 (0.69-1.55) | 0.94 (0.62-1.44) | 0.96 (0.64-1.44) |
| Non-HDL-C (≥130 vs. <130 mg/dL) | **1.84 (1.21-2.79)** | **1.69 (1.10-2.59)** | **1.67 (1.11-2.52)** |
| Triglycerides (≥150 vs. <150 mg/dL) | 1.31 (0.88-1.95) | 1.25 (0.82-1.91) | 1.20 (0.80-1.79) |
| Dyslipidemia | 1.27 (0.78-2.05) | 1.22 (0.74-2.02) | 1.19 (0.73-1.94) |
| **Plaque Progression** |  |  |  |
| Total cholesterol (≥200 vs. <200 mg/dL) | **1.83 (1.25-2.67)** | **1.79 (1.22-2.63)** | **1.69 (1.16-2.47)** |
| LDL-C (≥100 vs. <100 mg/dL) | 1.20 (0.83-1.74) | 1.25 (0.86-1.83) | 1.20 (0.83-1.74) |
| LDL-C (≥160 vs. <160 mg/dL) | **2.93 (1.51-5.70)** | **3.03 (1.53-5.98)** | **2.76 (1.43-5.33)** |
| LDL-C (mg/dL, continuous) | **1.009 (1.003-1.015)** | **1.009 (1.003-1.015)** | **1.008 (1.002-1.014)** |
| HDL-C (<40 vs. ≥40 mg/dL for men and <50 vs. ≥50 mg/dL for women) | 0.95 (0.65-1.39) | 0.87 (0.59-1.29) | 0.88 (0.61-1.29) |
| Non-HDL-C (≥130 vs. <130 mg/dL) | **1.57 (1.08-2.31)** | **1.53 (1.04-2.26)** | **1.47 (1.01-2.14)** |
| Triglycerides (≥150 vs. <150 mg/dL) | 1.31 (0.88-1.95) | 1.09 (0.74-1.61) | 1.03 (0.71-1.50) |
| Dyslipidemia | 1.23 (0.78-1.93) | 1.24 (0.77-1.99) | 1.16 (0.73-1.82) |
| **Incident Cardiovascular Disease** |  |  |  |
| Total cholesterol (≥200 vs. <200 mg/dL) | 1.27 (0.86-1.88) | 1.19 (0.80-1.78) | 1.20 (0.81-1.78) |
| LDL-C (≥100 vs. <100 mg/dL) | 0.94 (0.66-1.35) | 0.93 (0.65-1.34) | 0.96 (0.67-1.36) |
| LDL-C (≥160 vs. <160 mg/dL) | **2.78 (1.42-5.43)** | **2.75 (1.40-5.41)** | **2.75 (1.43-5.30)** |
| HDL-C (<40 vs. ≥40 mg/dL for men and <50 vs. ≥50 mg/dL for women) | 1.01 (0.77-1.58) | 1.03 (0.71-1.50) | 1.01 (0.71-1.44) |
| Non-HDL-C (≥130 vs. <130 mg/dL) | 1.18 (0.82-1.71) | 1.12 (0.76-1.63) | 1.14 (0.79-1.65) |
| Triglycerides (≥150 vs. <150 mg/dL) | 1.34 (0.94-1.91) | 1.21 (0.83-1.76) | 1.36 (0.96-1.94) |
| Dyslipidemia | 1.22 (0.86-1.72) | 1.17 (0.76-1.81) | 1.16 (0.77-1.77) |

\* During a median of 19 years of follow-up for CVD outcomes and 5 years of follow-up to identify plaque or plaque progression
†Multivariable models are adjusted for age, gender, current smoking, waist circumference, hypertension, diabetes mellitus, and
albuminuria. Reduced models are adjusted for age and diabetes.

### Association between dyslipidemia and clinical CVD

During a median follow-up of 18.5 years, 130 (9.2% of the 1,416 who were CVD free at baseline) participants developed fatal or nonfatal CVD events. Of these 130, 109 (83.8%) were MI and other CHD (such as undergoing percutaneous transluminal coronary angioplasty or coronary artery bypass grafting), 9 (6.9%) were stroke, 7 (5.4%) were CHF, and 5 (3.8%) were other fatal CVD; 10% (13/130) of all initial events were fatal. The five other fatal CVD events included 3 cases of cardiomyopathy and 2 cases of arrhythmia. From 2001 to 2020, the incidence rate of CVD among the study population was 5.4 per 1,000 person-years. Participants who were older at baseline had a significantly higher incidence of CVD than those who were younger. In addition, we observed a significantly higher CVD incidence in participants with increased waist circumference, hypertension, diabetes, metabolic syndrome, TC≥200 mg/dL, LDL-C≥160 mg/dL, non-HDL-C≥130 mg/dL, or TG≥150 mg/dL compared to those without these conditions (Figure 2). In the multivariable analysis, the risk of clinical CVD was higher in American Indians with versus without LDL-C ≥ 160 mg/dL, while controlling for covariates (HR = 2.75, 95% CI = 1.43-5.30, Table 4).

## Discussion

This is the first study to describe dyslipidemia among American Indian adolescents and young adults, a population with high prevalence of modifiable CVD risk factors. Dyslipidemia is important because it is a modifiable risk factor that has not been well assessed or controlled. We report that 70.6% of young American Indians had any type of dyslipidemia and 1.8% were taking lipid lowering medications during the baseline examination, 2001-2003. The use of lipid lowering medication remained low at the follow-up examination 2006-2009, with 8% taking medication. Although dyslipidemia is related to age, we demonstrated that every age group has an elevated prevalence of dyslipidemia with the prevalence being 55.2%, 73.6%, and 78.0% for participants 15-19, 20-29, and 30-39 years old, respectively. These estimates are substantially higher than that of the entire U.S. population where approximately 25% of adolsecents^14^ and 30% of young adults are expected to have dyslipidemia.^15^ Furthermore, approximately 2.8% had LDL-C ≥ 160 mg/dL, which is higher than the recommended threshold for medical along with lifestyle intervention in those under 40 years old.^9^ On an individual level 2.8% may not be alarming; however, on the population level 2.8% of very high LDL-C in people under 40 years old has tremendous clinical and public health significance. Despite the high baseline prevalence and the recommended thresholds for intervention, none of the participants who were under 20 years old were taking lipid lowering medication at baseline or 6-8 years later at follow-up. In American Indians with diabetes, aggressive lipid (LDL-C≤70 mg/dL) combined with blood pressure control demonstrated an improvement in carotid IMT, a measure of atherosclerosis, compared with standard control where IMT worsened.^10^ Since this population has an elevated prevalence of diabetes, this further demonstrates the need for lipid screening and control.^13,45^

The results for this younger American Indian population are consistent with previous reports where high LDL-C^5^ was associated with coronary heart disease (CHD) and stroke in American Indians who were 45-74 at baseline.^1,6^ Similarly, among adolescents and young adults, we report that clinical CVD was associated LDL-C ≥ 160 mg/dL independent of other CVD risk factors; however, we did not observe associations between CVD and TC, HDL-C, or TG. This is likely due to a limited number of incident clinical CVD events caused by participants being early in the disease process.^13^ Furthermore, we were able to establish an independent association between dyslipidemia measures (total cholesterol, LDL-C, non-HDL-C, triglycerides) and incident plaque, an early pathogenic CVD marker.^13^ We also observed independent associations between abnormal cholesterol levels and plaque progression. This indicates that dyslipidemia is not only associated with the development of plaque but also the worsening of plaque. These observations further illustrate the need for early screening and intervention of abnormal lipid levels in this population but also continued treatment to prevent clinical CVD.^10^

Regarding HDL-C, the results for this younger population are slightly different than analyses including older American Indians where low HDL-C was an independent risk factor of incident CVD.^46^ We observed no significant association between low levels of HDL-C and incident plaque, plaque progression, or clinical CVD. However, insulin plays an important role in lipid metabolism and therefore increased levels of TG in combination with decreased levels of HDL-C are highly prevalent in people with diabetes.^47^ This is likely due to the over production of very low-density lipoproteins (VLDL), which alters the composition of HDL and precedes the diagnosis of type 2 diabetes by several years.^1,48^ This is further demonstrated by the diabetes prevalence in older versus younger participants, which were 40% and 8.2%, respectively.^1^ Therefore, it is likely that this younger population will develop HDL dyslipidemia if the prevalence of diabetes increases.

A limitation of this study is that the results may only be generalizable to other populations with high burden of cardiometabolic risk factors such as hyperglycemia, dyslipidemia, and abdominal obesity. However, as 47 million U.S. residents are living with at least one cardiometabolic disorder,^49^ we hope our findings may attract more attention and resources to address dyslipidemia screening and management in adolescents and young adults living with high burden of cardiometabolic disorders. Another limitation is that we did not have enough statistical power to evaluate CVD subcategories, such as CHD and stroke. Although we had adequate power to detect an association between LDL-C and combined events, future investigation is needed to study associations between dyslipidemia and subtypes of CVD events. Furthermore, the overall definitions of abnormal cholesterol levels were largely developed with data from non-Native older adults and may not be appropriate for younger American Indians.^9^ Also, the Friedwald equation may underestimate LDL-C when triglycerides are higher.^22,50^ However, we directly measured LDL-C when triglycerides were very high (> 400 mg/dL) and the Friedwald equation is a validated method that used extensively in clinical practice.^50^ If anything, the definition of dyslipidemia based on older adults and the Friedwald equation for LDL-C calculation likely underestimate the prevalence and association observed through these analyses. Despite these potential limitations, the data we present clearly establish the associations between dyslipidemia, subclinical atherosclerosis, and clinical CVD in younger American Indians in the Strong Heart Family Study.

In conclusion, we demonstrated that elevated levels of TC, LDL-C, and non-HDL-C were independently associated with incident plaque and plaque progression, indicators of subclinical CVD. Furthermore, elevated LDL-C was associated with clinical CVD in a population of adolescents and young adults where we would not expect to observe such clinical manifestations. Due to all these factors, young American Indians and other young populations with high burden of cardiometabolic disorders are likely to benefit significantly from lipid screening and cholesterol health education, lifestyle modification, and medical interventions at an early age.

## Data Availability

Data are available for approval from the Strong Heart Study Publications and Presentations Committee, the Strong Heart Study Steering Committee, the participating Tribes, and the Indian Health Service.

https://strongheartstudy.org/Research/Study-Data-and-Study-Samples

https://strongheartstudy.org/

## Acknowledgements and Sources of Funding

The SHS has been funded in whole or in part with federal funds from the National Heart, Lung, and Blood Institute, National Institute of Health, Department of Health and Human Services, under contract numbers 75N92019D00027, 75N92019D00028, 75N92019D00029, & 75N92019D00030 and research grants: R01HL109315, R01HL109301, R01HL109284, R01HL109282, and R01HL109319 and cooperative agreements: U01HL41642, U01HL41652, U01HL41654, U01HL65520, and U01HL65521. The content is solely the responsibility of the authors and does not necessarily represent official views of the National Institutes of Health or the Indian Health Service.

